# HIGH AND UNSUSTAINABLE COST OF CONTROLLING THE DENGUE VECTOR AEDES AEGYPTI IN THE PERUVIAN AMAZON

**DOI:** 10.1101/2023.12.19.23300246

**Authors:** Salomón Durand, Arles Paredes, Carlos Pacheco, Ray Fernandez, Jose Herrera, Cesar Cabezas

## Abstract

**Objectives:** Vector control is the method of prevention and control of dengue outbreaks used in Peru. With the objective of calculating the costs incurred in vector control, an estimation of the costs of *Aedes aegypti* control of the Regional Health Directorate of Loreto, during the execution of the regional plan for surveillance and control of *A. aegypti* was carried out.

**Materials and methods:** Documentation was reviewed, and interviews were conducted with stakeholders about the costs of *A. aegypti* control interventions in its adult and larval stages, during 2017 and 2018.

**Results:** It was found that the costs incurred in the control of the dengue vector in the Loreto Region in the two years studied amount to: 3,807,858 and 4,066,380 soles during 2017 and 2018 respectively, 1’175,264 and 1’1210,232 dollars at the exchange rate of 2017 and 2018, however, the effect of the control activities is short-lived.

**Conclusions:** The high cost involved in vector control with the methods currently used and the short duration of its effect makes it unsustainable so studies should be conducted to find other more efficient methods of dengue control.

## INTRODUCTION

Since its reintroduction in 1990, dengue has become the most important vector-borne disease in Peru^(1)^. Dengue is caused by an arbovirus, DENV, and transmitted by a mosquito, *Aedes aegypti* (*A. aegypti*), and *Aedes albopictus* (however, the presence of albopictus has not yet been reported in Peru), which causes outbreaks during the rainy season or when the temperature rises, which can cause health services to collapse, as occurred in Iquitos in 2011, after the entry of the Asian/American genotype of DENV-2^(2)^.

Since the first report of its reintroduction to Peru in 1984, *A. aegypti* has spread from the Amazon to almost the entire country. The mobility of people due to trade, migration or other reasons facilitates the involuntary transport of vector eggs in containers, infesting new localities^(3)^. 21 of Peru’s 24 departments have the presence of *A. aegypti* and in 20 there is dengue transmission^(3)^. In the urban centers of the Amazon and the coast, this disease is prevalent and outbreaks are also currently being reported in small rural communities, which aggravates the problem^(1)^.

In the 1950s, the elimination of this vector in the country was possible within the framework of urban yellow fever eradication campaigns^(4)^, but, at present, despite multiple efforts, this vector has not been controlled in a sustained manner^(1)^.

The strategy used in Peru and other endemic countries to reduce dengue transmission is the control of its vector, *A. aegypti*, in its different stages. The main approach is the use of larvicides or inhibitors of its development in water collection containers for drinking water, the elimination of waste and the use of insecticides for adult mosquitoes inside and outside homes. This strategy has been used for decades and has been evaluated for its effectiveness and sustainability^(5)^. In Peru, few evaluation studies or studies testing new control methodologies have been published^(6)^. Likewise, the few published cost evaluation studies conducted in Peru do not describe the cost of vector control in a disaggregated manner^(7,8)^.

Surveillance and vector control activities for *A. aegypti* in Peru are financed by the state through regional governments, municipalities and the Ministry of Health^(9)^. The Regional Health Directorate of Loreto (DIRESA Loreto), through the Environmental Health Directorate (DESA), conducts annual interventions to control *A. aegypti* through two strategies: larval control, through entomological surveillance (aedic surveys), control with the application of larvicide growth inhibitors, both through the inspection of high-risk households throughout the territory of the Loreto region; and adult mosquito control through space fogging with portable equipment (portable aerosol generators and thermal foggers) in accordance with the regulations of the Ministry of Health^(9)^.

This study was carried out to determine the costs incurred in the control of *A. aegypti* in a region of the Peruvian Amazon.

## MATERIALS AND METHODS

The study was conducted in the Loreto region, which has a population of 1’077,831 inhabitants, of which approximately 500,000 live in the city of Iquitos, a city located on the banks of the Amazon River. Iquitos has poor basic sanitation and water distribution only for a few hours, so families store water in containers such as cylinders, buckets and pots, which become potential breeding grounds for the vector, and high temperatures and constant rains favor infestation of homes^(6)^.

This study is descriptive and was conducted retrospectively; it was planned as a partial economic evaluation of intervention costs. Costs were estimated from the perspective of the public health system^(10)^, considering that the Ministry of Health (MINSA), within the framework of the technical standard for surveillance and vector control of dengue in Peru^(9)^, is the financier through the Regional Health Directorate of Loreto (DIRESA). All direct and indirect costs of the vector control program were included.

Vector control interventions for dengue control carried out from January 2017 to December 2018 were analyzed; for which the costs of interventions for both larval control, as well as adult mosquito control, carried out according to regulations^(9)^, including scheduled vector control or in response to outbreaks, were reviewed.

Data were collected using forms designed for this purpose, which compiled the information according to the classification of expenditures or costs (direct sanitary, indirect and unit costs). All the proposed dengue prevention, surveillance and control plans were requested from the DIRESA’s Environmental Health Directorate; then documentation was requested for all the vector control interventions carried out during the 2 years of the study, in compliance with the established plans and interventions in response to outbreaks; in addition to payment slips, receipts, payments and were also compared with the purchases and services entered into the Integrated Administrative Management System (SIGA). Interviews were also conducted with the professional responsible for vector control and biologists directly involved in *A. aegypti* surveillance and control activities of the Environmental Health Directorate; logistics and human resources personnel were interviewed to acquire the costs of equipment and vehicle purchases; in addition to salaries of personnel working directly or indirectly in vector control interventions.

To estimate the cost of inputs, such as the insecticides malathion 57% Emulsion Concentrate (EC) (for adult mosquito control) and pyriproxyfen 0.5% (for control in the immature stages), the database of MINSA’s National Center for the Supply of Strategic Health Resources (CENARES) was reviewed, which specifies the unit cost and annual acquisitions.

Costs were classified as proposed by Drummond^(11)^, classifying costs into health and non-health costs; health costs are the costs related to the health intervention and its subsequent evolution and treatment, which are assumed by the health system. They include the time of health professionals; the price of supplies, personal protective equipment and sanitary products used, among others.

The direct health costs of the implementation of each intervention were collected according to each strategy carried out during the fieldwork, either from the annual periodic programming or in response to outbreaks. The plans and reports of each intervention carried out (larval control, adult vector control) were evaluated. Direct health costs were divided into the costs of personnel directly involved in vector control interventions (brigade chief, fumigation operators, registrars, drivers, megaphone advertising personnel, etc.); support personnel also directly involved (vector control center director, administrative technicians, biologists, etc.); materials and supplies; and equipment and vehicles.

Indirect health costs were calculated for the salaries of personnel working in the Environmental Health Directorate who participate indirectly in vector control interventions, costs of renting premises, mobilities, office expenses, security, etc. Only the cost of water and electricity of the main facility was considered and not the other attached facilities as they were considered not significant. For the cost of equipment and goods, straight-line depreciation was used at a rate of 10%, considering the calculated useful life of the equipment.

The information collected was analyzed using Microsoft Excel ® and SPSS 22 (IBM ®). Costs were reported in soles and converted to U.S. dollars using the average exchange rate for each year of the study, which was 3.4 soles. The study was approved by the research committee of the Regional Health Directorate of Loreto.

## RESULTS

It was found that for the year 2017, the cost of dengue vector control was S/. 3,807,858.73, while in 2018 it was S/. 4,066,380.25. Larval control in general caused higher cost with S/. 2,562,881.50 in 2017 and S/. 2,239,406.50 in 2018. (see table 01).

**Table 01:** Summary of total costs incurred in vector control interventions of Aedes aegypti, Regional Health Directorate Loreto, 2017 - 2018.

| **COSTS OF DENGUE VECTOR CONTROL -**<br>**DIRESA** | **2018** | **2017** |
| --- | --- | --- |
| Adult mosquito control - Spatial nebulization<br>with portable equipment | S/ 1,709,110.25 | S/ 1,108,071.73 |
| Larval control | S/ 2,239,406.50 | S/ 2,562,881.50 |
| Aedic surveys | S/ 117,863.50 | S/ 134,888.50 |
| **TOTAL** | **S/. 4,066,380.25** | **S/. 3,807,858.73** |

DIRESA= Regional Health Directorate Loreto

Table 02 shows the costs of the interventions of space fogging campaigns with portable equipment to control the vector in its adult stage (mosquito), the costs are divided into direct health costs and indirect health costs. For 2017, only 2 interventions or space fogging campaigns with portable equipment were carried out in the prioritized sectors of the city of Iquitos. Regarding direct sanitary costs, this was divided into personnel who participate directly in the campaigns, with a total cost of S/. 599,040.00; support personnel, who work in the vector control office with a total cost of S/. 57,140.00; then there are the inputs, protection, and logistical materials with a total cost of S/. 301,393.90; among these is the adulticide insecticide (malathion at 57% EC) with an annual cost of S/. 28,560.50 and fuel was the highest cost in this item, finally, there are the costs of equipment, machinery, and transportation; with an annual cost of S/. 40,020.83. The total direct sanitary cost was S/. 997,594.73 per year, while the total indirect sanitary cost was S/. 108,460.00.

**Table 02:** Annual cost of A. aegypti control campaigns, adult mosquito: direct and indirect health costs. DIRESA LORETO, 2017 - 2018.

| **DIRECT SANITARY COSTS** |  |  |  |
| --- | --- | --- | --- |
| **Personal de intervención fumigación** | **2017** | **2018** | **TOTAL** |
|  | **Costo** | **Costo** |  |
| General Supervisors - Biologists | 35,100.00 | 38,610.00 |  |
| Brigade Chiefs | 39,780.00 | 42,120.00 |  |
| Space Nebulization Operators | 397,800.00 | 421,200.00 |  |
| Others | 126,360.00 | 136,890.00 |  |
| **Total** | **599,040.00** | **638,820.00** | **1,237,860.00** |
| **Support Personnel** |  |  |  |
| Vector Control Surveillance Officer | 6,720.00 | 10,080.00 |  |
| Biologists | 11,200.00 | 16,800.00 |  |
| Administrative Technician | 3,240.00 | 4,860.00 |  |
| Others | 35,980.00 | 53,970.00 |  |
| **Total** | **57,140.00** | **85,710.00** | **142,850.00** |
| **Supplies and Materials** |  |  |  |
| Petroleum | 109,250.00 | 257,266.50 |  |
| 84 Octane Gasoline | 75,000.00 | 23,940.00 |  |
| Insecticide: 57% Malathion | 28,560.50 | 44,454.00 |  |
| Others | 88,583.40 | 429,268.50 |  |
| **Total** | **301,393.90** | **754,929.00** | **1,056,322.90** |
| **Equipment, Machinery and Transportation** |  |  |  |
| Moto pulverizadora | 20,187.50 | 30,281.25 |  |
| Moto - Vans | 2,666.67 | 4,150.00 |  |
| Others | 17,166.67 | 25,750.00 |  |
| **Total** | **40,020.83** | **60,181.25** |  |
| **Total, direct sanitary costs** | **997,594.73** | **1,539,640.25** | **S/ 2,437,032.90** |
| **INDIRECT SANITARY COSTS** |  |  |  |
| **Personal de sede central DESA** | **2017** | **2018** | **TOTAL** |
|  | **Costo** | **Costo** |  |
| Director | 2,240.00 | 5,040.00 |  |
| Administrator | 1,280.00 | 2,880.00 |  |
| Others | 2,160.00 | 3,240.00 |  |
| **Total** | **5,680.00** | **11,160.00** | **16,840.00** |
| **Office and Miscellaneous Expenses** |  |  |  |
| Office Materials and Equipment | 2,500.00 | 3,750.00 |  |
| Transport rental (colectivos o buses) | 96,000.00 | 144,000.00 |  |
| Others | 4,280.00 | 10,560.00 |  |
| **Total** | **102,780.00** | **158,310.00** | **261,090.00** |
| **Total, indirect sanitary costs** | **108,460.00** | **169,470.00** | **S/ 277,930.00** |
| **TOTAL COST OF MOSQUITO CONTROL** | **S/. 1,106,054.73** | **S/. 1,709,110.25** | **S/. 2,714,962.90** |

DESA = Environmental Sanitation Directorate
DIRESA=Loreto Regional Health Directorate

During 2018, 3 space fogging campaigns were carried out with portable equipment in prioritized sectors of the city of Iquitos. Regarding direct health costs, annual cost for personnel directly involved in the campaigns was S/. 638,820.00; annual cost for support personnel was S/. 85,710.00; for supplies, protection and logistical materials, the annual cost was S/. 754,929.00; there being a large difference in this item compared to the previous year, because more campaigns were carried out, in addition, in 2018 the purchase of 90 octane gasoline and a greater amount of adulticide insecticide (malathion) was carried out with an annual cost of S/. 44,454.00. Equipment and transportation costs only reached an annual cost of S/. 60,181.25. and, finally, there is the annual indirect sanitary cost of S/. 169,470.00. 169,470.00 (see table 02).

As for larval vector control, this is carried out in a programmed manner, in five campaigns or intervention cycles of 2 months duration each cycle. These activities include surveillance activities such as aedic surveys, home inspections and the use of larval growth controllers or inhibitors, as well as the elimination of breeding sites and guidance to households on prevention. There were 130 home inspectors distributed in different establishments in the region, 108 in the city of Iquitos and 22 in the periphery; these inspectors carried out the aedic survey and the application of pyriproxyfen 0.5% for larval control.

Table 3 shows the costs of the intervention of inspection of homes at risk of dengue transmission and the application of aedic surveys, this is divided into direct and indirect health costs; as for direct health costs, this was divided into personnel directly involved in the inspection of homes and aedic survey, with a total cost for the year 2017 of S/. 2,025,600.00; support staff working in the vector control office with a total cost of S/. 83,160.00; then there are inputs, protection, and logistic materials with a total cost of S/. 549,185.00; within these is the granulated insecticide Piriproxifen 0.5% with a cost of S/. 375,900.00; and finally, there are other expenses such as training S/. 7,225.00; and aedic survey S/. 134,888.50.

**Table 03:** Annual cost of home inspections and medical survey: direct and indirect health costs. DIRESA, 2017 – 2018.

| **COSTS OF DENGUE VECTOR CONTROL -DIRESA LORETO** |  |  |  |
| --- | --- | --- | --- |
| **EXPENSES GENERATED IN VECTOR CONTROL - LARVARIO - YEAR 2017-2018** |  |  |  |
| **DIRECT HEALTH COSTS** |  |  |  |
| **Senior Professional Staff** | **2017** | **2018** | **TOTAL** |
|  | **Costo** | **Costo** |  |
| Brigade Chiefs | 580,800.00 | 660,000.00 | 1,240,800.00 |
| Housing Inspectors | 1,372,560.00 | 1,231,200.00 | 2,603,760.00 |
| Aedic Survey | 72,240.00 | 64,800.00 | 137,040.00 |
| **TOTAL** | **2,025,600.00** | **1,956,000.00** | **3,981,600.00** |
| **Support Personnel** |  |  |  |
| Biologists | 60,000.00 | 60,000.00 |  |
| Director of the Vector Control Center | 13,440.00 | 13,440.00 |  |
| Administrative Technician | 9,720.00 | 9,720.00 |  |
| **Total** | **83,160.00** | **83,160.00** | **166,320.00** |
| **Supplies and Material** |  |  |  |
| Piripoxifen 0.5% | 375,900.00 | 105,000.00 |  |
| Gloves | 39,000.00 | 39,000.00 |  |
| Backpack | 32,500.00 | 32,500.00 |  |
| Tshirts | 7,800.00 | 7,800.00 |  |
| Others | 31,336.50 | 42,751.50 |  |
| Materials for Aedic Survey | 62,648.50 | 51,433.50 |  |
| **Total** | **549,185.00** | **278,485.00** | **827,670.00** |
| **Other Expenses** |  |  |  |
| Training | 7,225.00 | 7,225.00 | 14,450.00 |
| Aedic Survey | 134,888.50 | 116,233.50 | 251,122.00 |
| **Total Direct Sanitary Costs** | **2,665,170.00** | **2,324,870.00** | **4,990,040.00** |
| **INDIRECT SANITARY COSTS** |  |  |  |
| **DESA Central Office Personnel** |  |  |  |
| Executive Director | 4,500.00 | 4,500.00 |  |
| Administrator | 3,500.00 | 3,500.00 |  |
| Others | 4,200.00 | 4,200.00 |  |
| **Total** | **12,200.00** | **12,200.00** | **24,400.00** |
| **Office and Miscellaneous Expenses** |  |  |  |
| Office Material and Equipment | 5,000.00 | 5,000.00 |  |
| Rental of main headquarters | 11,000.00 | 11,000.00 |  |
| Others | 4,200.00 | 4,200.00 |  |
| **Total** | **20,200.00** | **20,200.00** | **40,400.00** |
| **Total Indirect Sanitary Costs** | **32,400.00** | **32,400.00** | **64,800.00** |
| **TOTAL COST LARVAL CONTROL** | **2,697,570.00** | **2,357,270.00** | **5,054,840.00** |

DESA = Environmental Sanitation Directorate DIRESA=Loreto Regional Health Directorate

In 2018, the cost in housing inspector personnel was S/. 1,956,000.00; the cost for support personnel was S/. 83,160.00; for supplies, personal protection and logistical materials the cost was S/. 278,485.00, within this cost is included the granulated insecticide Piriproxifen at 0. Finally, there are other expenses such as training for inspectors at a cost of S/. 7,225.00; and an aedic survey at a cost of S/. 116,233.50. As for indirect sanitary costs, for both 2017 and 2018 it was a total of S/. 32,400.00 per year.

## DISCUSSION

It was found in this study that the total annual cost of *A. aegypti* control in Loreto was S/. 3,807,858.73 for 2017 (1,175,264 USD) and S/. 4,066,380.25 for 2018 (1,210,232 USD).

An economic evaluation of the vector control interventions for *A. aegypti*, carried out during 2017 and 2018; by the Regional Directorate of Health Loreto; these interventions were basically of 2 types, larval control, which includes preventive type activities such as aédic survey, inspection of homes and the use of larval growth regulators, and control of the adult vector by spatial fogging with light or portable equipment in which the insecticide malathion is applied, insecticide with an efficiency greater than 97% mortality at that time, (Pacheco C., personal communication).

Vector control activities in Peru are carried out in accordance with MINSA regulations^(9)^. Vector control is carried out in scenarios II and III (presence of the vector with sporadic cases and presence of the vector in outbreaks). In scenario II, the objective is to reduce the risk of dengue transmission and is aimed at controlling Aedes in its larval stage. In scenario III, the objective is to rapidly control transmission and Aedes control methods are applied in both the larval and adult stages.

The costs found are high compared to those reported in other countries, considering that Loreto has about one million inhabitants; however, countries where dengue is transmitted, like Peru, dedicate significant resources to vector control^(12-15)^. There are few studies on costs in Peru and the few reports do not describe in detail the expenses incurred in vector control. In Piura in 2002, an expense of S/. 64,260 in 50 days was reported for vector control of a dengue outbreak in the town of Sachura^(16)^. Salmon-Mulanovich et al. in 2014 calculated the cost of families in the care of cases in a dengue outbreak in an Amazon region finding that the cost for each case was on average 105.3 dollars^(7)^. Hans-Christian Stahl et al. estimated the cost of the 2011 outbreak in Peru at US$ 4.5 million, of which 16% corresponded to vector control (US$ 738,701)^(8)^.

The vector control methods used would be effective in reducing vector density^(17)^; programmed vector control (larval control, insecticides, biologicals, etc.) would cost less than vector control in response to outbreaks. In Peru, the effectiveness of vector control has been evaluated. Stoddard et al, in a study that recorded data for a decade in the city of Iquitos, concluded that there is evidence of the impact of vector control of adults on dengue transmission if it is applied early in outbreaks^(18)^. On the other hand, in the city of Iquitos, Reiner et al. elaborated a model based on the density of *A. aegypti* adults, collected over several years, which shows that depending on the coverage of houses achieved in spatial fogging, a reduction in the density of female adults responsible for transmission can be achieved between 67 and 43% if 100 or 50% coverage of intervened houses is achieved, respectively^(19)^.

Although it is true that the methods used would be effective in reducing vector density, their effect is short-lived; according to Pontes et al. larval control would have a persistence of only 2 months in the best of cases^(20)^. Control of adults with ultra-low volume insecticide applications has an immediate effect on the mosquito population that would last only 1 day according to Koenraadt^(21)^. In this sense, to maintain the density of *A. aegypti* at low levels to reduce dengue transmission, it is necessary to maintain larval control cycles and carry out adult control, all of this at a high cost, not achieving prolonged or definitive control with the techniques currently used. Likewise, we must consider that, in outbreaks with a large, affected population, it is sometimes necessary to increase the frequency of interventions for adult control; in 2011, more than 10 interventions for adults were performed in one year due to the magnitude of the outbreak (Rodriguez H., personal communication).

In addition, the effect of interventions based on insecticides would diminish over time due to the emergence of vector resistance, having to replace them with molecules of higher cost and toxicity.

There would be other conditions where it is possible to intervene, for example, in Iquitos, as in other cities in the country, there is a water supply of less than 5 hours a day, which forces the population to store water in all types of containers that become breeding grounds. of vector^(3)^. Likewise, the accumulation of unusable waste and frequent rains create the conditions for the growth of the vector.

Vector control of Aedes aegypti does not prevent the seasonal increase in dengue cases and outbreaks that occur in Loreto, as shown in Figure 1.

**Graph 01:**
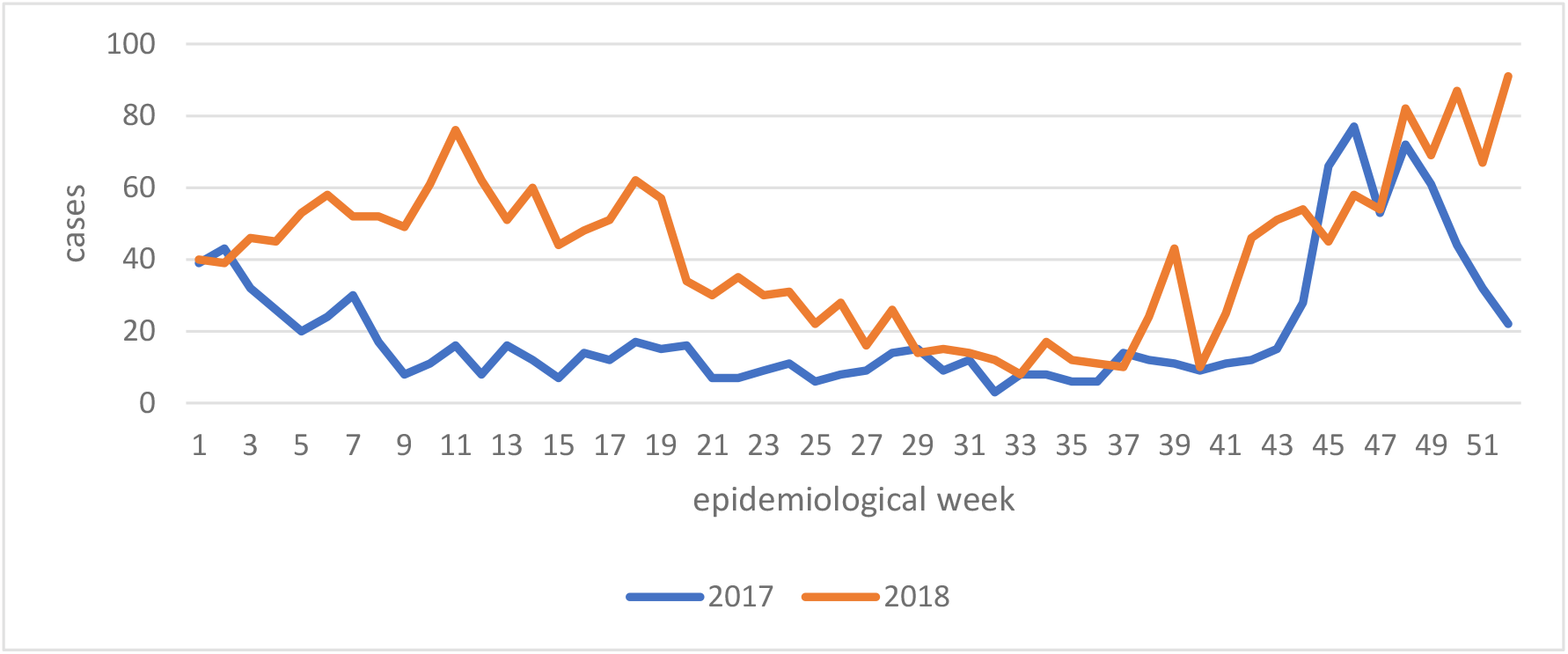
Dengue cases reported in the Loreto region by epidemiological week during the study period (2017 – 2018) Source: Report from the Epidemiology Directorate of Loreto Regional Health Directorate

A recent phenomenon observed is the invasion of A. aegypti from rural communities, probably due to the adoption of customs that facilitate the development of the vector, which would increase the costs of control^(22)^.

One limitation of this study was that the analysis of adult mosquito control (spatial fogging with portable equipment) was based mainly on data from the city of Iquitos, because data from interventions in the provinces are difficult to quantify and because of underreporting of activities. Likewise, the differentiated costs of water and electricity and the water and electricity costs of other rented premises were not considered in this analysis because they were not considered significant in comparison to other costs. On the other hand, the evaluation was carried out in years with moderate reporting of cases and not in a year with intense transmission such as the one that occurred in 2011 in Loreto.

Taking into account that this economic evaluation was partial^(23)^ and did not estimate the benefits or results on the health of the population, it is possible, however, that this study could serve as a baseline for subsequent studies to evaluate the impact, considering that the poor quality of the data and the existing underreporting in endemic countries make it difficult to analyze the impact of vector control^(24)^.

In conclusion, the costs incurred in dengue vector control in the Loreto Region in the two years studied exceed US$1 million per year, and while it is true that interventions have an immediate effect in reducing transmission, this effect is short-lived, creating a dependence on the use of insecticides that are costly to apply.

Basing dengue prevention on vector control with the methods currently used is unsustainable in a country like Peru with multiple public health problems and the risk of epidemics of other communicable diseases. For these reasons, it would be necessary to evaluate other methods of prevention and control of dengue in Peru.

## Data Availability

All data produced in the present work are contained in the manuscript

## Notes

**Conflict of Interest:** None of the authors declare conflicts of interest.

### Competing Interest Statement

The authors have declared no competing interest.

### Funding Statement

This study did not receive any funding

